# A Reassessment of Sodium Correction Rates and Hospital Length of Stay Accounting for Admission Diagnosis

**DOI:** 10.1101/2024.03.08.24303993

**Authors:** Eric Raphael Gottlieb, Leo Anthony Celi

**Author notes:** Corresponding Author: Eric R. Gottlieb, MD, MS Cambridge Health Alliance 1493 Cambridge St. Cambridge, MA 02139.

## Abstract

**Background:** Slow correction of severe hyponatremia has been historically recommended due to the risk of rare but catastrophic neurologic events with rapid correction. A recent study challenging this paradigm reported that rapid correction is associated with shorter hospital length of stay, but that study did not control for admission diagnosis. The objective of this study was to determine whether rapid correction is associated with shorter length of stay when controlling for admission diagnosis.

**Methods:** This retrospective cohort study is based on the fourth edition of the Medical Information Mart for Intensive Care, MIMIC-IV, a deidentified, publicly available clinical research database which includes admissions from 2008-2019. Patients were identified who presented to the hospital with initial sodium <120 mEq/L and were categorized according to total sodium correction achieved in the first day (<6 mEq/L; 6-10 mEq/L; >10 mEq/L). Linear regression was used to assess for an association between correction rate and hospital length of stay, and to determine if this association was significant when controlling for admission diagnosis classifications based on diagnosis related groups (DRGs).

**Results:** There were 419 patients with severe hyponatremia (<120 mEq/L) included in this study, of whom 374 survived to discharge. Median [IQR] hospital length of stay was 6 [4, 11] days. In a univariable linear regression, there was a trend towards a significant association between the highest rate of correction (>10 mEq/L) and shorter length of stay, as compared with a moderate rate of correction (coef. -2.764, 95% CI [-5.791, 0.263], p=0.073), but the association was not significant when controlling for admission diagnosis group (coef. -1.561, 95% CI [-4.398, 1.276], p=0.280). There was a significant association in the survivor subset (coef. -3.455, 95% CI [-6.668, -0.242], p=0.035), but it was also not significant when controlling for admission diagnosis group (coef. -2.200, 95% CI [-5.144, 0.743], p=0.142).

**Conclusions:** Rapid correction is not associated with shorter length of stay when controlling for admission diagnosis, suggesting that the disease state confounds this association. Findings from prior and future studies reporting this association should not drive clinical decision making if the confounding effect of hospital admission diagnosis and competing risk of death are not fully accounted for.

## Introduction

Hyponatremia is a common primary or secondary hospital admission diagnosis. For patients with chronic severe hyponatremia, rapid correction of sodium levels is commonly thought to increase the risk of osmotic demyelination syndrome (ODS), previously known as central pontine myelinolysis (CPM), a potentially catastrophic neurologic event.^1^ Therefore, longstanding recommendations have stressed slow correction of chronic hyponatremia.^2^

However, the risks and benefits of slow correction have recently been questioned, given the possibility of adverse effects of delaying normalization of sodium levels, combined with the rarity of ODS, as well as debate about the connection between sodium correction rates and ODS.^3^ In light of this, several recent studies have re-examined outcomes associated with rapid correction of severe hyponatremia.^4,5^ In particular, Seethapathy et al. (2023) showed that faster correction was associated with shorter hospital length of stay and lower mortality.

While these and other findings could be practice-changing, it is important to recognize that the studies did not directly account for admission diagnosis, leaving open the potential for significant confounding. This is a known concern with numerous prior studies, leading the authors of one recent review article to question whether reported associations between correction rates and mortality are “causality or epiphenomenon.”^6^

If these associations are confounded or confused by competing risk, such as when the outcome of length of stay is defined as “time from admission to discharge or death” as per Seethapathy et al. (2023),^4^ acting on them clinically would not be beneficial and might even be dangerous for some patients. Given the publication of these recent studies that clinicians may cite to support rapid correction strategies for their patients, it is important to examine these factors critically.

The Medical Information Mart for Intensive Care (MIMIC), a deidentified predominately critical care research database developed and maintained through a longstanding collaboration between the Beth Israel Deaconess Medical Center (BIDMC) in Boston, MA and the Massachusetts Institute of Technology, can be used to assess the impact of admission diagnosis by providing information on Diagnosis Related Groups (DRGs). DRGs classify admissions according to the primary problem addressed and may be tied to hospital reimbursement, but may or may not overlap with chronic comorbidities that are more frequently used in retrospective analyses.^7,8^

In this study, we use these data to determine how admission diagnosis modifies the association between sodium correction rate and hospital length of stay in an overall cohort and a surviving subset of patients with severe hyponatremia, defined as serum sodium <120 mEq/L.^3^

## Methods

### Research Ethics

This retrospective cohort study is based on the Medical Information Mart for Intensive Care, fourth iteration (MIMIC-IV), a primarily critical care research database with data from 2008-2019.^9^ MIMIC was approved for research by the institutional review boards of BIDMC (2001-P-001699/14) and MIT (0403000206) without a requirement for individual patient informed consent because data are deidentified and publicly available.

### Cohort Selection

Data were obtained via the Google BigQuery (Alphabet Inc.) cloud platform using RStudio Version 2024.04.02 (Posit Software, PBVC) with the R 4.4.0 programming language (R Foundation for Statistical Computing).

Hospital admissions were identified where the first sodium value was less than 120 mEq/L. Patients were excluded if they were less than 18 years of age and/or were admitted for less than 24 hours. For each patient, only the last hospital admission was included to maximally assess the competing risk of mortality. This study followed the STROBE guidelines for observational studies.^10^

### Sodium Value Determination

The initial sodium value used for each patient was the first charted value for the admission. The final sodium value was the value from closest to 24 hours after the initial value, but was limited to values between 20 and 28 hours later. This method was used rather than approximating a 24-hour value to better reflect real-world management. The total sodium correction during this period was calculated as the difference between the final value and the initial value and was classified for analysis as less than 6 mEq/L, 6-10 mEq/L, or greater than 10 mEq/L, as was done previously.^4^

### Disease and Organ System Classifications

DRGs for admissions were classified by disease state and/or organ system, based on manual review of the data set to identify common categories, as well as disease states commonly associated with hyponatremia. The final categories chosen were: cardiopulmonary, digestive, hematology/oncology, infection, liver, toxic/metabolic, neurologic/psychiatric, orthopedic, renal/urologic, and other diagnosis. If a patient fit both a disease state and an organ system (e.g. infection and liver), the patient was classified into the non-organ-based disease state group (infection). Patients categorized primarily by the electrolyte disorder were included in the toxic/metabolic category. The final classifications for all DRGs used in this study are provided in e-Table 1.

**Table 1:** Characteristics of patients by correction rate.

| Variable | Overall | <6 mEq/L | 6-10 mEq/L | >10 mEq/L |
| --- | --- | --- | --- | --- |
| N | 419 | 168 | 164 | 87 |
| Age (years) | 68.0 [57.0, 80.0] | 70.0 [60.0, 82.0] | 68.0 [56.0, 80.0] | 63.0 [55.0, 77.5] |
| Male (%) | 190 (45.3) | 78 (46.4) | 80 (48.8) | 32 (36.8) |
| Race/Ethnicity (%) |  |  |  |  |
| Asian | 33 (7.9) | 14 (8.3) | 9 (5.5) | 10 (11.5) |
| Black | 27 (6.4) | 10 (6.0) | 9 (5.5) | 8 (9.2) |
| Hispanic | 18 (4.3) | 10 (6.0) | 3 (1.8) | 5 (5.7) |
| White | 300 (71.6) | 126 (75.0) | 120 (73.2) | 54 (62.1) |
| Other/Unknown Ethnicity | 41 (9.8) | 8 (4.8) | 23 (14.0) | 10 (11.5) |
| Diagnosis |  |  |  |  |
| Toxic/Metabolic | 193 (46.1) | 76 (45.2) | 72 (43.9) | 45 (51.7) |
| Renal/Urologic | 27 (6.4) | 5 (3.0) | 14 (8.5) | 8 (9.2) |
| Cardiopulmonary | 59 (14.1) | 28 (16.7) | 23 (14.0) | 8 (9.2) |
| Digestive | 9 (2.1) | 3 (1.8) | 4 (2.4) | 2 (2.3) |
| Hematology/Oncology | 13 (3.1) | 8 (4.8) | 3 (1.8) | 2 (2.3) |
| Infection | 39 (9.3) | 13 (7.7) | 14 (8.5) | 12 (13.8) |
| Liver | 35 (8.4) | 16 (9.5) | 17 (10.4) | 2 (2.3) |
| Neurologic/Psychiatric | 13 (3.1) | 3 (1.8) | 7 (4.3) | 3 (3.4) |
| Orthopedic | 12 (2.9) | 6 (3.6) | 5 (3.0) | 1 (1.1) |
| Other | 19 (4.5) | 10 (6.0) | 5 (3.0) | 4 (4.6) |
| Hypertonic Saline (%) | 92 (22.0) | 31 (18.5) | 45 (27.4) | 16 (18.4) |
| Initial Na | 117.0 [115.0, 119.0] | 117.0 [115.0, 118.0] | 118.0 [115.0, 119.0] | 117.0 [113.5, 119.0] |
| Final Na | 123.0 [119.0, 127.0] | 119.0 [117.0, 121.0] | 125.0 [123.0, 126.0] | 130.0 [128.0, 133.0] |
| Length of Stay (days) | 6.4 [4.1, 10.7] | 7.3 [5.0, 11.1] | 6.7 [4.2, 11.9] | 5.0 [3.1, 7.7] |
| Deaths (%) | 45 (10.7) | 19 (11.3) | 20 (12.2) | 6 (6.9) |

### Determination of Elixhauser Scores

International classification of diseases (ICD) revision 10 codes were extracted to evaluate acute diagnoses and chronic comorbidities collectively. For each hospital admission, the Elixhauser comorbidity score was calculated^11^ using the R ‘comorbidity’^12^ package. This score aggregates ICD codes into a set of 31 binary diagnostic categories.^13,14^ For each DRG category specified in this study, the proportion of patients who carried ICD codes consistent with the DRG-based diagnosis category was also determined, as shown in e-Table 2.

**Table 2:** Proportions of patients in each diagnosis group and mortality, length of stay, and correction rate proportions for that diagnosis.

| Organ/Disease Category | Proportion of Patients with Admission Diagnosis (%) | Mortality (%) | Length of Stay (days)<sup>1</sup> | Proportion <6 mEq/L (%) | Proportion 6-10 mEq/L (%) | Proportion >10 mEq/L (%) |
| --- | --- | --- | --- | --- | --- | --- |
| Toxic/Metabolic | 46 | 4 | 5 [3, 7] | 39 | 37 | 23 |
| Cardiopulmonary | 14 | 15 | 9 [5, 16] | 47 | 39 | 14 |
| Infection | 9 | 28 | 8 [5, 13] | 33 | 36 | 31 |
| Liver | 8 | 29 | 16 [8, 25] | 46 | 49 | 6 |
| Renal/Urologic | 6 | 0 | 6 [5, 9] | 19 | 52 | 30 |
| Other | 5 | 5 | 9 [6, 18] | 53 | 26 | 21 |
| Hematology/Oncology | 3 | 15 | 6 [4, 10] | 62 | 23 | 15 |
| Neurologic/Psychiatric | 3 | 31 | 7 [4, 13] | 23 | 54 | 23 |
| Orthopedic | 3 | 0 | 8 [4, 16] | 50 | 42 | 8 |
| Digestive | 2 | 11 | 13 [10, 17] | 33 | 44 | 22 |

<sup>1</sup>Median [IQR]

### Statistical Analysis

Univariable linear regression analysis was used to assess for an association between correction rate category in the first day (<6 mEq/L; 6-10 mEq/L; >10 mEq/L) and hospital length of stay, and multivariable regression analysis with the addition of a multi-level variable for diagnosis category was used to determine if the observed association persisted when controlling for diagnosis group. The reference correction rate was 6-10 mEq/L and the reference diagnosis in the multivariable models was toxic/metabolic. This analysis was repeated with the exclusion of patients for whom that hospitalization ended in death, to assess the impact of the competing risk of death on the association, if any, between sodium correction rate and hospital length of stay. For all analyses, a p-value of <0.05 was considered statistically significant.

## Results

There were 419 patients included in this study, of whom 374 survived to discharge, giving a mortality rate of 11%. Overall cohort characteristics are shown in Table 1. Patients were 55% female and had a median [interquartile range, IQR] age of 68 [57, 80] years. Patients had a median [IQR] initial sodium value of 117 [115, 119] mEq/L and final sodium value of 123 [119, 127] mEq/L. Median [IQR] initial and final sodium values for patients in each correction rate category are shown in Table 1. Proportions of patients with corrections of <6 mEq/L, 6-10 mEq/L, and >10 mEq/L were 40%, 39%, and 21%, respectively. As shown in Table 2, these proportions differed widely by organ system/disease state.

Patients undergoing rapid (>10 mEq/L) initial correction in the first day vs. moderate (6- 10 mEq/L) and slow correction (<6 mEq/L) were in general younger, less likely to be male, less likely to be White, more likely to have toxic/metabolic and infection admission diagnoses, and less likely to have liver and cardiopulmonary diagnoses. Overall, 77% of patients had an Elixhauser diagnosis that matched their DRG category, but also with substantial variation by DRG category (e-Table 2).

The median [IQR] hospital length of stay was 6 [4, 11] days. Hospital lengths of stay stratified by survival for each admission diagnosis are shown in Figure 1. Median [IQR] lengths of stay and mortality by correction rate are shown in Table 1.

**Figure 1:**
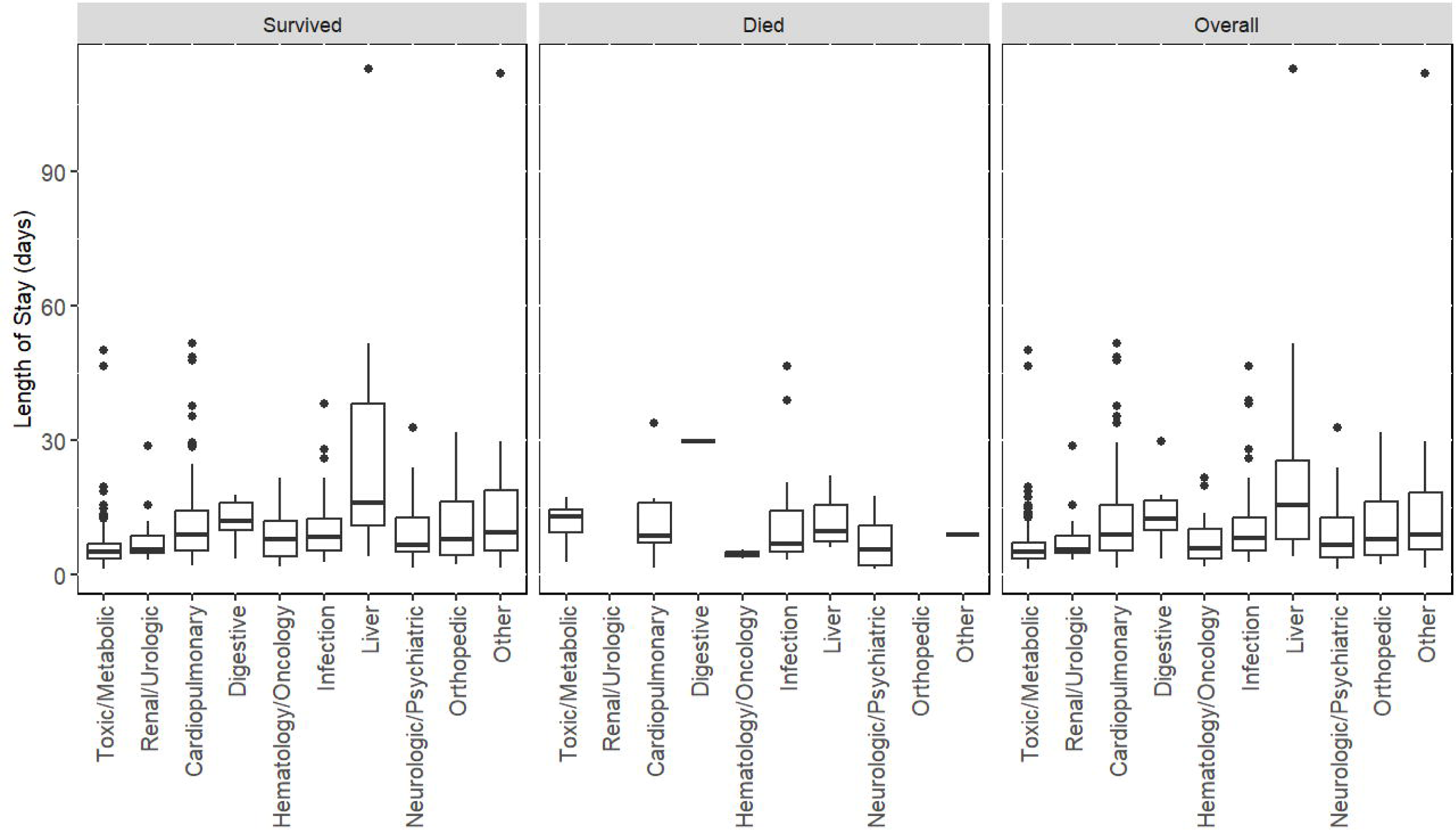
Hospital length of stay by diagnosis in patients who survived and died, and in the overall cohort.

As shown in Table 3, in a univariable linear regression, there was a trend towards a significant association between the highest rate of correction (>10 mEq/L) and shorter length of stay, as compared with a moderate rate of correction (6-10 mEq/L, coef. -2.764, 95% CI [-5.791, 0.263], p=0.073), but the association was not significant when controlling for admission diagnosis group (coef. -1.561, 95% CI [-4.398, 1.276], p=0.280). This association was significant in a univariable analysis for the majority subset of patients who survived the hospitalization (coef. -3.455, 95% CI [-6.668, -0.242], p=0.035), but it was also not significant when controlling for admission diagnosis group (coef. -2.200, 95% CI [-5.144, 0.743], p=0.142). There was no difference in length of stay between patients who underwent slow (<6 mEq/L) and moderate correction in either the full cohort (coef. 1.600, 95% CI [-0.905, 4.105], p=0.210) or in survivors (coef. 1.690, 95% CI [-1.013, 4.394], p=0.220) in a univariable analysis.

**Table 3:** Linear regression models of associations between correction rate and length of stay, controlling for organ system/disease state.

|  | All | +Organ/Disease | Survived | +Organ/Disease |
| --- | --- | --- | --- | --- |
| (Intercept) | 10.007<br>[8.225,11.789]<br>p<0.001 | 6.022<br>[3.993,8.051]<br>p<0.001 | 9.921<br>[7.993,11.849]<br>p<0.001 | 5.955<br>[3.857,8.052]<br>p<0.001 |
| Rate <6 | 1.600<br>[-0.905,4.105]<br>p=0.210 | 1.468<br>[-0.883,3.820]<br>p=0.220 | 1.690<br>[-1.013,4.394]<br>p=0.220 | 1.556<br>[-0.920,4.032]<br>p=0.217 |
| Rate >10 | -2.764<br>[-5.791,0.263]<br>p=0.073 | -1.561<br>[-4.398,1.276]<br>p=0.280 | -3.455<br>[-6.668,-0.242]<br>p=0.035 | -2.200<br>[-5.144,0.743]<br>p=0.142 |
| Cardiopulmonary |  | 6.687<br>[3.528,9.845]<br>p<0.001 |  | 6.988<br>[3.635,10.342]<br>p<0.001 |
| Digestive |  | 7.951<br>[0.735,15.168]<br>p=0.031 |  | 6.168<br>[-1.405,13.741]<br>p=0.110 |
| Hematology/Oncology |  | 1.794<br>[-4.286,7.875]<br>p=0.562 |  | 2.786<br>[-3.731,9.304]<br>p=0.401 |
| Infection |  | 5.632<br>[1.912,9.352]<br>p=0.003 |  | 5.067<br>[0.809,9.324]<br>p=0.020 |
| Liver |  | 15.153<br>[11.239,19.066]<br>p<0.001 |  | 19.266<br>[14.776,23.755]<br>p<0.001 |
| Neurologic/Psychiatric |  | 3.982<br>[-2.092,10.056]<br>p=0.198 |  | 4.866<br>[-2.308,12.040]<br>p=0.183 |
| Orthopedic |  | 4.641<br>[-1.666,10.947]<br>p=0.149 |  | 4.717<br>[-1.545,10.979]<br>p=0.139 |
| Renal/Urologic |  | 1.710<br>[-2.660,6.080]<br>p=0.442 |  | 1.950<br>[-2.395,6.295]<br>p=0.378 |
| Other |  | 10.227<br>[5.132,15.322]<br>p<0.001 |  | 10.877<br>[5.694,16.061]<br>p<0.001 |
| N | 419 | 419 | 374 | 374 |

## Discussion

In this study, we leverage high-resolution clinical and administrative data to assess how controlling for hospital diagnosis categories modifies associations between early sodium correction rate and hospital length of stay in patients with severe hyponatremia. Following a recent study showing that a correction of >10 mEq/L in the first 24 hours of hospitalization was associated with shorter length of stay,^4^ we report a consistent strong trend towards such an association, but when controlling for admission diagnosis categories, the association is clearly not significant.

When limiting the analysis to patients who survived the hospitalization, the univariable association becomes stronger despite a somewhat smaller cohort, but it is also abrogated by controlling for diagnosis category. The stronger association in the surviving cohort reflects how the competing risk of death confuses assessment of hospital length of stay, but the confounding by diagnosis remains profound. Prior studies that have neither adequately controlled for admission diagnosis, nor managed the effect of hospital mortality, such as when length of stay is defined as “time from admission to discharge or death,”^4^ are thus vulnerable to missing important confounding and competing factors. As a result, these findings may inappropriately be used by clinicians as evidence to support rapid correction strategies.

Management of severe hyponatremia is a controversial subject in nephrology and critical care, and it can have a significant impact on the hospital course. It can also have major medicolegal implications, as adverse outcomes may be attributed to overly rapid correction.^2^ Slow correction is recommended to reduce the still-disputed risk of ODS, but adding to the rarity of this outcome, data showing improved outcomes with faster correction, including more easily observable proxy outcomes such as hospital length of stay, could shift this balance.^4,15,16^

Confounding by diagnosis has presented a critical shortcoming in prior analyses.^6^ Conceptually, it can be understood by considering that patients with certain diagnoses, such as decompensated liver disease, may present in a critically ill state that portends a prolonged hospitalization, and it may not be safe or physiologically possible to rapidly correct their sodium values.^17^ Meanwhile patients presenting with more readily reversible conditions that do not portend a prolonged admission may also be clinically better candidates for rapid sodium correction. However, it is not clear that correcting sodium more rapidly in a patient with decompensated liver disease, if possible, would reduce length of stay, and doing so could lead to other complications that could even increase length of stay.

The use of diagnosis related groups (DRGs), a standard classification of hospital admission diagnoses which we categorized according to organ systems or general disease states, is a novel strategy to reassess previous findings while addressing confounding by diagnosis. For this analysis, DRGs are preferable to other more commonly used sources of information about diagnosis, such as ICD codes, which may not always distinguish between acute presentations and underlying chronic conditions.^18^ As we show, using common comorbidity burden indices such as the Elixhauser score may also capture some admission diagnoses better than others, making them poor substitutes for direct classification of admission diagnosis and potentially introducing further bias.

It is important to reiterate that while our study does not support an overall shorter length of stay with faster correction, it does not show that rapid correction is detrimental either.

However, it supports the need for more nuanced consideration of hospital diagnosis in future studies and at the bedside. It also stresses that it may be inadvisable to manage sodium correction according to one-size-fits-all guidelines. We find this to be true with regard to diagnosis, but also across providers and institutions, speaking to the need for local and continuous validation of these findings.

Strengths of this study include the use of DRG categories to more precisely identify the reason for admission. We provide all applicable DRGs and our associated classifications in e- Table 1, allowing for scrutiny and reproducibility. We also clearly remove the competing risk of death in our final analysis. Lastly, this study was based on high-resolution data, and we assessed correction rates in a way that mimicked real-world conditions rather than artificial averages.^4^

Limitations are that our study only included patients with severe hyponatremia, defined as initial sodium <120 mEq/L. Disease state classifications are subjective, and changing these classifications could theoretically affect the results. Given that MIMIC-IV is limited to inpatient laboratory data, and as is often the case in real-life practice, we did not have prior laboratory values to prove that hyponatremia was chronic, which is generally a necessary prerequisite to recommend slow correction. However, the prior study also did not have this historical data, and it is not clear that known acute hyponatremia was excluded, as patients had to have a sodium level less than 120 mEq/L, but it was not necessarily the first value, as we required in the present study.^4^ Our cohort was also smaller than some prior work on this topic supporting faster sodium correction, but it was large enough to demonstrate significance in the cohort of patients who survived the hospitalization, and given that it was based on only two multi-level variables, it substantially exceeded the previously determined number of subjects per variable for linear regression.^19^ The sample size also was not large enough to assess mortality or very rare neurologic events, which would be better suited to a case-control study design.^4,20^

## Conclusion

When controlling for admission diagnosis categories, there was not a significant association between relatively rapid sodium correction rate and shorter hospital length of stay. Our findings do not support faster sodium correction as a strategy to reduce hospital length of stay in a diverse population of patients admitted to the hospital with severe hyponatremia.

Further research is needed to determine subsets of patients likely to benefit from or be harmed by rapid correction.

## Data Sharing Statement

MIMIC is publicly available with training in human subjects research and application. Statistical code is available upon request.

## Data Availability

All data used in this manuscript are available online at https://mimic.mit.edu/.

