## Supplemental Tables for "A Reassessment of Sodium Correction Rates and Hospital Length of Stay Accounting for Admission Diagnosis"

e-Table 1: DRG Diagnoses and Associated Categories

| Description | Disease Group | N |
| --- | --- | --- |
| MISC DISORDERS OF NUTRITION,METABOLISM,FLUIDS/ELECTROLYTES W/O MCC | Toxic/Metabolic | 52 |
| ENDOCRINE DISORDERS W CC | Toxic/Metabolic | 34 |
| ENDOCRINE DISORDERS W MCC | Toxic/Metabolic | 32 |
| MISC DISORDERS OF NUTRITION,METABOLISM,FLUIDS/ELECTROLYTES W MCC | Toxic/Metabolic | 31 |
| ENDOCRINE DISORDERS W/O CC/MCC | Toxic/Metabolic | 20 |
| HEART FAILURE & SHOCK W MCC | Cardiopulmonary | 14 |
| RENAL FAILURE W CC | Renal/Urologic | 12 |
| NUTRITIONAL & MISC METABOLIC DISORDERS W/O MCC | Toxic/Metabolic | 11 |
| DISORDERS OF LIVER EXCEPT MALIG,CIRR,ALC HEPA W MCC | Liver | 10 |
| CIRRHOSIS & ALCOHOLIC HEPATITIS W MCC | Liver | 8 |
| NUTRITIONAL & MISC METABOLIC DISORDERS W MCC | Toxic/Metabolic | 8 |
| RENAL FAILURE W MCC | Renal/Urologic | 8 |
| LIVER TRANSPLANT W MCC OR INTESTINAL TRANSPLANT | Liver | 6 |
| SEPTICEMIA OR SEVERE SEPSIS W/O MV 96+ HOURS W MCC | Infection | 6 |
| ACUTE MYOCARDIAL INFARCTION, DISCHARGED ALIVE W MCC | Cardiopulmonary | 5 |
| SEPTICEMIA OR SEVERE SEPSIS W/O MV >96 HOURS W MCC | Infection | 5 |
| CIRRHOSIS & ALCOHOLIC HEPATITIS W CC | Liver | 4 |
| HEART FAILURE & SHOCK W CC | Cardiopulmonary | 4 |
| INFECTIOUS & PARASITIC DISEASES W O.R. PROCEDURE W MCC | Infection | 4 |
| EXTENSIVE O.R. PROCEDURE UNRELATED TO PRINCIPAL DIAGNOSIS W MCC | Other | 3 |
| HEART FAILURE & SHOCK W MCC OR PERIPHERAL EXTRACORPOREAL MEMBRANE OXYGEN | Cardiopulmonary | 3 |
| INTRACRANIAL HEMORRHAGE OR CEREBRAL INFARCTION W MCC | Neurologic/Psychiatric | 3 |
| MAJOR GASTROINTESTINAL DISORDERS & PERITONEAL INFECTIONS W MCC | Infection | 3 |
| MAJOR SMALL & LARGE BOWEL PROCEDURES W MCC | Digestive | 3 |
| MALIGNANCY OF HEPATOBILIARY SYSTEM OR PANCREAS W MCC | Hematology/Oncology | 3 |
| OTHER RESP SYSTEM O.R. PROCEDURES W MCC | Cardiopulmonary | 3 |
| RESPIRATORY SYSTEM DIAGNOSIS W VENTILATOR SUPPORT 96+ HOURS | Cardiopulmonary | 3 |
| SEPTICEMIA OR SEVERE SEPSIS W MV 96+ HOURS | Infection | 3 |
| AMPUTATION FOR CIRC SYS DISORDERS EXC UPPER LIMB & TOE W MCC | Orthopedic | 2 |
| CARDIAC ARRHYTHMIA & CONDUCTION DISORDERS W MCC | Cardiopulmonary | 2 |
| CARDIAC VALVE & OTH MAJ CARDIOTHORACIC PROC W CARD CATH W MCC | Cardiopulmonary | 2 |
| CESAREAN SECTION W CC/MCC | Other | 2 |
| COMPLICATIONS OF TREATMENT W MCC | Other | 2 |
| DIABETES W CC | Toxic/Metabolic | 2 |
| DIGESTIVE MALIGNANCY W CC | Hematology/Oncology | 2 |
| ECMO OR TRACH W MV 96+ HRS OR PDX EXC FACE, MOUTH & NECK W MAJ O.R. | Cardiopulmonary | 2 |
| ECMO OR TRACH W MV >96 HRS OR PDX EXC FACE, MOUTH & NECK W MAJ O.R. | Cardiopulmonary | 2 |
| G.I. HEMORRHAGE W MCC | Digestive | 2 |
| HEPATOBILIARY DIAGNOSTIC PROCEDURES W MCC | Liver | 2 |
| INFLAMMATION OF THE MALE REPRODUCTIVE SYSTEM W/O MCC | Other | 2 |
| INTRACRANIAL HEMORRHAGE OR CEREBRAL INFARCTION W CC | Neurologic/Psychiatric | 2 |
| KIDNEY & URINARY TRACT INFECTIONS W/O MCC | Infection | 2 |
| OTHER RESP SYSTEM O.R. PROCEDURES W CC | Cardiopulmonary | 2 |
| PERC CARDIOVASC PROC W NON-DRUG-ELUTING STENT W MCC OR 4+ VES/STENTS | Cardiopulmonary | 2 |
| PERIPHERAL VASCULAR DISORDERS W MCC | Other | 2 |
| RESPIRATORY INFECTIONS & INFLAMMATIONS W CC | Infection | 2 |
| SEPTICEMIA OR SEVERE SEPSIS W MV >96 HOURS | Infection | 2 |
| SIMPLE PNEUMONIA & PLEURISY W MCC | Infection | 2 |
| TRACH W MV 96+ HRS OR PDX EXC FACE, MOUTH & NECK W/O MAJ O.R. | Cardiopulmonary | 2 |
| TRAUMATIC STUPOR & COMA, COMA <1 HR W CC | Neurologic/Psychiatric | 2 |
| ACUTE MYOCARDIAL INFARCTION, EXPIRED W MCC | Cardiopulmonary | 1 |
| ADRENAL & PITUITARY PROCEDURES W CC/MCC | Toxic/Metabolic | 1 |
| ALCOHOL/DRUG ABUSE OR DEPENDENCE W/O REHABILITATION THERAPY W MCC | Toxic/Metabolic | 1 |
| AMPUTATION FOR MUSCULOSKELETAL SYS & CONN TISSUE DIS W CC | Orthopedic | 1 |
| BIOPSIES OF MUSCULOSKELETAL SYSTEM & CONNECTIVE TISSUE W CC | Orthopedic | 1 |
| BRONCHITIS & ASTHMA W CC/MCC | Cardiopulmonary | 1 |
| CELLULITIS W/O MCC | Infection | 1 |
| CHRONIC OBSTRUCTIVE PULMONARY DISEASE W CC | Cardiopulmonary | 1 |
| CHRONIC OBSTRUCTIVE PULMONARY DISEASE W MCC | Cardiopulmonary | 1 |
| CIRCULATORY DISORDERS EXCEPT AMI, W CARD CATH W MCC | Cardiopulmonary | 1 |
| COAGULATION DISORDERS | Hematology/Oncology | 1 |
| CRANIOTOMY & ENDOVASCULAR INTRACRANIAL PROCEDURES W MCC | Neurologic/Psychiatric | 1 |
| DIABETES W MCC | Toxic/Metabolic | 1 |
| DIGESTIVE MALIGNANCY W MCC | Hematology/Oncology | 1 |
| DISORDERS OF THE BILIARY TRACT W CC | Liver | 1 |
| DISORDERS OF THE BILIARY TRACT W MCC | Liver | 1 |
| ESOPHAGITIS, GASTROENT & MISC DIGEST DISORDERS W/O MCC | Digestive | 1 |
| EXTENSIVE O.R. PROCEDURE UNRELATED TO PRINCIPAL DIAGNOSIS W CC | Other | 1 |
| FOOT PROCEDURES W CC | Orthopedic | 1 |
| FX, SPRN, STRN & DISL EXCEPT FEMUR, HIP, PELVIS & THIGH W/O MCC | Orthopedic | 1 |
| HEPATOBILIARY DIAGNOSTIC PROCEDURES W CC | Liver | 1 |
| HIP & FEMUR PROCEDURES EXCEPT MAJOR JOINT W MCC | Orthopedic | 1 |
| HIV W MAJOR RELATED CONDITION W MCC | Infection | 1 |
| HIV W OR W/O OTHER RELATED CONDITION | Infection | 1 |
| INFECTIONS, FEMALE REPRODUCTIVE SYSTEM W CC | Infection | 1 |
| INFLAMMATORY BOWEL DISEASE W MCC | Digestive | 1 |
| KIDNEY & URETER PROCEDURES FOR NON-NEOPLASM W MCC | Renal/Urologic | 1 |
| KIDNEY & URINARY TRACT SIGNS & SYMPTOMS W/O MCC | Renal/Urologic | 1 |
| LOCAL EXCISION & REMOVAL INT FIX DEVICES EXC HIP & FEMUR W CC | Orthopedic | 1 |
| LYMPHOMA & NON-ACUTE LEUKEMIA W MCC | Hematology/Oncology | 1 |
| MAJOR CARDIOVASC PROCEDURES W MCC | Cardiopulmonary | 1 |
| MAJOR CARDIOVASC PROCEDURES W MCC OR THORACIC AORTIC ANEURYSM REPAIR | Cardiopulmonary | 1 |
| MAJOR HEMATOL/IMMUN DIAG EXC SICKLE CELL CRISIS & COAGUL W CC | Hematology/Oncology | 1 |
| MAJOR JOINT & LIMB REATTACHMENT PROC OF UPPER EXTREMITY W CC/MCC | Orthopedic | 1 |
| MALIGNANCY, FEMALE REPRODUCTIVE SYSTEM W MCC | Hematology/Oncology | 1 |
| MEDICAL BACK PROBLEMS W/O MCC | Orthopedic | 1 |
| MYELOPROLIF DISORD OR POORLY DIFF NEOPL W MAJ O.R. PROC W MCC | Hematology/Oncology | 1 |
| NON-EXTENSIVE O.R. PROC UNRELATED TO PRINCIPAL DIAGNOSIS W MCC | Other | 1 |
| NONSPECIFIC CEREBROVASCULAR DISORDERS W MCC | Neurologic/Psychiatric | 1 |
| NONTRAUMATIC STUPOR & COMA W MCC | Neurologic/Psychiatric | 1 |
| OTHER ANTEPARTUM DIAGNOSES W MEDICAL COMPLICATIONS | Other | 1 |
| OTHER CIRCULATORY SYSTEM DIAGNOSES W MCC | Cardiopulmonary | 1 |
| OTHER DIGESTIVE SYSTEM DIAGNOSES W MCC | Digestive | 1 |
| OTHER DIGESTIVE SYSTEM O.R. PROCEDURES W MCC | Digestive | 1 |
| OTHER DISORDERS OF NERVOUS SYSTEM W MCC | Neurologic/Psychiatric | 1 |
| OTHER EAR, NOSE, MOUTH & THROAT O.R. PROCEDURES W CC/MCC | Other | 1 |
| OTHER HEPATOBILIARY OR PANCREAS O.R. PROCEDURES W MCC | Liver | 1 |
| OTHER INFECTIOUS & PARASITIC DISEASES DIAGNOSES W CC | Infection | 1 |
| OTHER KIDNEY & URINARY TRACT DIAGNOSES W CC | Renal/Urologic | 1 |
| OTHER KIDNEY & URINARY TRACT PROCEDURES W CC | Renal/Urologic | 1 |
| OTHER KIDNEY & URINARY TRACT PROCEDURES W MCC | Renal/Urologic | 1 |
| OTHER MUSCULOSKELET SYS & CONN TISS O.R. PROC W CC | Orthopedic | 1 |
| OTHER O.R. PROCEDURES FOR MULTIPLE SIGNIFICANT TRAUMA W MCC | Other | 1 |
| OTHER VASCULAR PROCEDURES W MCC | Other | 1 |
| PANCREAS, LIVER & SHUNT PROCEDURES W MCC | Liver | 1 |
| PERC CARDIOVASC PROC W DRUG-ELUTING STENT W MCC OR 4+ VESSELS/STENTS | Cardiopulmonary | 1 |
| PERMANENT CARDIAC PACEMAKER IMPLANT W CC | Cardiopulmonary | 1 |
| PERMANENT CARDIAC PACEMAKER IMPLANT W MCC | Cardiopulmonary | 1 |
| POSTOPERATIVE & POST-TRAUMATIC INFECTIONS W MCC | Infection | 1 |
| PULMONARY EMBOLISM W MCC | Cardiopulmonary | 1 |
| RESPIRATORY INFECTIONS & INFLAMMATIONS W MCC | Infection | 1 |
| RESPIRATORY NEOPLASMS W CC | Hematology/Oncology | 1 |
| RESPIRATORY NEOPLASMS W MCC | Hematology/Oncology | 1 |
| SEIZURES W MCC | Neurologic/Psychiatric | 1 |
| SEPTIC ARTHRITIS W MCC | Infection | 1 |
| SEPTICEMIA W/O MV 96+ HOURS W MCC | Infection | 1 |
| SIMPLE PNEUMONIA & PLEURISY W CC | Infection | 1 |
| SKIN GRAFT &/OR DEBRID EXC FOR SKIN ULCER OR CELLULITIS W CC | Other | 1 |
| SKIN GRAFTS & WOUND DEBRID FOR ENDOC, NUTRIT & METAB DIS W CC | Other | 1 |
| TRACH W MV >96 HRS OR PDX EXC FACE, MOUTH & NECK W/O MAJ O.R. | Cardiopulmonary | 1 |
| TRANSURETHRAL PROSTATECTOMY W CC/MCC | Renal/Urologic | 1 |
| TRAUMATIC STUPOR & COMA, COMA <1 HR W MCC | Neurologic/Psychiatric | 1 |
| UPPER LIMB & TOE AMPUTATION FOR CIRC SYSTEM DISORDERS W CC | Orthopedic | 1 |
| URINARY STONES W/O ESW LITHOTRIPSY W/O MCC | Renal/Urologic | 1 |

e-Table 2: Proportion of patients with corresponding Elixhauser diagnosis by disease group

| Disease Group | % Corresponding |
| --- | --- |
| Toxic/Metabolic^1^ | 100 |
| Renal/Urologic^2^ | 52 |
| Cardiopulmonary^3^ | 98 |
| Digestive^4^ | 0 |
| Hematology/Oncology^5^ | 92 |
| Infection^6^ | 5 |
| Liver^7^ | 91 |
| Neurologic/Psychiatric^8^ | 62 |
| Orthopedic^9^ | 17 |
| Other | 0 |
| ^1^Corresponding Elixhauser diagnoses: fluid and electrolyte disorders, alcohol abuse, drug abuse, complicated diabetes, uncomplicated diabetes, hypothyroidism, obesity, weight loss | |
| ^2^Corresponding Elixhauser diagnosis: renal failure | |
| ^3^Corresponding Elixhauser diagnoses: congestive heart failure, cardiac arrhythmia, valvular disease, pulmonary circulation disorders, chronic pulmonary disease, complicated hypertension, uncomplicated hypertension | |
| ^4^Corresponding Elixhauser diagnosis: peptic ulcer disease | |
| ^5^Corresponding Elixhauser diagnosis: deficiency anemia, solid tumor without metastasis, metastatic cancer, blood loss anemia, coagulopathy, lymphoma | |
| ^6^Corresponding Elixhauser diagnosis: HIV/AIDS | |
| ^7^Corresponding Elixhauser diagnosis: liver disease | |
| ^8^Corresponding Elixhauser diagnoses: other neurologic disorders, psychosis, depression, paralysis | |
| ^9^Corresponding Elixhauser diagnosis: rheumatoid arthritis | |
